# A genome-resolved view of the wastewater RNA virome

**DOI:** 10.64898/2026.05.19.26353600

**Authors:** Rose S. Kantor, Migun Shakya, Nelson Ruth, Jason A. Rothman, Clayton Rushford, Devon A. Gregory, Aidan Epstein, Jeff T. Kaufman, Jonathan E. Allen, Patrick S. G. Chain, David H. O’Connor, Marc C. Johnson

**Affiliations:** Physical and Life Sciences Directorate, Lawrence Livermore National Laboratory, Livermore, CA; Bioscience Division, Los Alamos National Laboratory, Los Alamos, NM; Department of Microbiology and Plant Pathology, University of California, Riverside, Riverside, CA; Department of Molecular Microbiology and Immunology, University of Missouri-School of Medicine, Columbia, MO; Global Security Computing Applications Division, Lawrence Livermore National Laboratory, Livermore, CA; SecureBio, Cambridge, MA; Department of Pathology and Laboratory Medicine, University of Wisconsin-Madison, Madison, WI

**Author notes:** These authors contributed equally (Rose S. Kantor, Migun Shakya).

## Abstract

Sequencing-based wastewater surveillance is emerging as an important tool in pathogen-agnostic threat detection, potentially enabling early identification before capture through clinical surveillance systems. However, virus sequences of human pathogens are typically low in abundance in wastewater while much of the data is unclassifiable at the read level. This presents a challenge because genomes may not assemble well for novel pathogens of interest, but read-based methods cannot currently separate novel from previously seen unclassified sequences. Using ultra-deep untargeted sequencing of the wastewater RNA virome performed by the CASPER consortium (321 samples), we constructed a wastewater virus genome database (“WVDB”) with the goal of expanding the set of available high-quality non-redundant reference genomes. The first version of this database contains 21,015 near-complete viral genomes, of which the majority are ssRNA bacteriophage (79%). We additionally recovered genomes for putative plant and vertebrate-infecting viruses, human enteric viruses, and viruses whose host could not be predicted. Fewer than 4000 genomes had matches in previously published virus genome databases, and WVDB captured around one fifth of the reads that could not be classified by Kraken2. Further expansion of WVDB will provide a comprehensive resource of RNA virus genomes for characterization of viral diversity and dynamics in wastewater across space and time.

## 1. Introduction

Sequencing-based wastewater biosurveillance has provided valuable information during the COVID-19 pandemic and beyond. Specifically, sequencing has been used to identify the circulation of known and novel SARS-CoV-2 variants ^1–4^, the occurrence of zoonotic influenza A subtypes ^5^, and the presence of measles in new locations ^6^. The principal methodological challenge in wastewater virus sequencing is that, even after physical concentration of viruses in wastewater (e.g. ultrafiltration, polyethylene glycol precipitation or skim milk flocculation), vertebrate-infecting RNA viruses are rare, while bacteriophage and plant viruses are common and abundant ^7,8^. Tiled amplicon or probe-capture hybridization methods can substantially increase the abundance of human-infecting viruses of public health interest ^9^, but depending on the targets, these panels may completely miss or underrepresent novel or emerging viral threats.

Ultra-deep untargeted sequencing is a complementary approach to increase sensitivity while remaining pathogen-agnostic. Yet untargeted sequencing presents a unique challenge - the recovery of a vast set of unknown sequences. Efforts to illuminate the wastewater virome have been pursued since at least the early 2010’s ^8,10–13^, but early work was limited to shallow sequencing due to the cost of the technology at the time ^14^. Recent untargeted efforts have explored the wastewater virome either deeply in a few locations ^15,16^, or globally with shallower sequencing ^17–19^. Some have successfully recovered complete genomes or tracked strains of human pathogens over space and time ^15,16^. Although valuable, these efforts have not provided publicly available high-quality genomes, making it difficult to draw comparisons between studies and to annotate unknown viral reads in new wastewater samples.

The CASPER project has produced the largest collection of ultra-deep untargeted wastewater sequencing data to-date, described in detail by Justen *et al.* (2026), Rushford *et al.* (2025), and Grimm *et al.* (2025)^14,20,21^. Given that this project targets ∼1 billion reads per sample and primarily focuses on human health objectives, most analyses of this data have thus far been read-based ^14,20^. That approach makes sense for human respiratory viruses, because of their low abundance and because reference databases include high-quality sequences for these viruses already. However, novel sequences of concern by definition cannot be detected via reference-based methods (or they may be incompletely detected). Here, *de novo* assembly approaches can help, either by directly recovering the genomes of concerning viruses, or indirectly by recovering genomes of novel benign viruses, thereby allowing annotation of previously unclassified reads within the dataset and removing them from consideration as threats.

Recent *de novo* assembly efforts utilizing publicly available metagenomic data have illustrated the benefits of an assembly-based approach to recover viral “dark matter” across a range of environments ^22,23^ including the human body ^24^ and gut ^25^, but relatively few metatranscriptomic datasets were available for inclusion, likely limiting the recovery of RNA viruses. Given the depth and breadth of raw wastewater sequencing in the CASPER effort and the deliberate bias toward RNA sequencing^14,20,21^, these datasets have the potential to yield additional viral genomes beyond those previously reported.

Here, we describe the construction of a wastewater virus genome database (WVDB) from a subset of 321 samples collected as part of the CASPER project. Our aims were to expand the availability of high-quality reference genomes for viruses in wastewater, describe the breadth of observed RNA viruses, and infer their distribution across geographical settings. This database and associated analyses are intended to grow as more CASPER data is incorporated.

## 2. Results

We compiled metagenomic sequencing data from untreated wastewater collected at 11 locations across six cities in the United States between 2023-2025 (**Figure 1A**). The dataset totaled 321 samples and included a one-year weekly time series from Columbia, Missouri, as well as samples from an airport (IL_Chicago_C). Samples were sequenced to a median depth of 1.6 billion reads (range 0.5 - 5.7; **Figure S1**). From the single assemblies of all metagenomes, we collected 195,826 near-complete (≥90% completeness estimated by checkV) viral genomes (**Figures 1B; S2A**). After clustering at the level of viral operational taxonomic units (vOTUs) and requiring two independent genome assemblies per vOTU to address potential misassemblies (**Figure S2B**), we recovered 21,015 unique, high-quality vOTU genomes (**Figure 1C**). These genomes and metadata, available at https://zenodo.org/records/20276352, are termed the Wastewater Virome Database (WVDB). A total of 21,343 vOTUs were not included in the final database because they were singletons and/or the possibility of misassemblies could not be ruled out (**Figure S2B**). In parallel, 1,458,420 RNA dependent RNA polymerase (RdRp) genes recovered from single assemblies were clustered to produce a database containing 199,032 RdRp representatives (**Figure S3A**).

**Figure 1.**
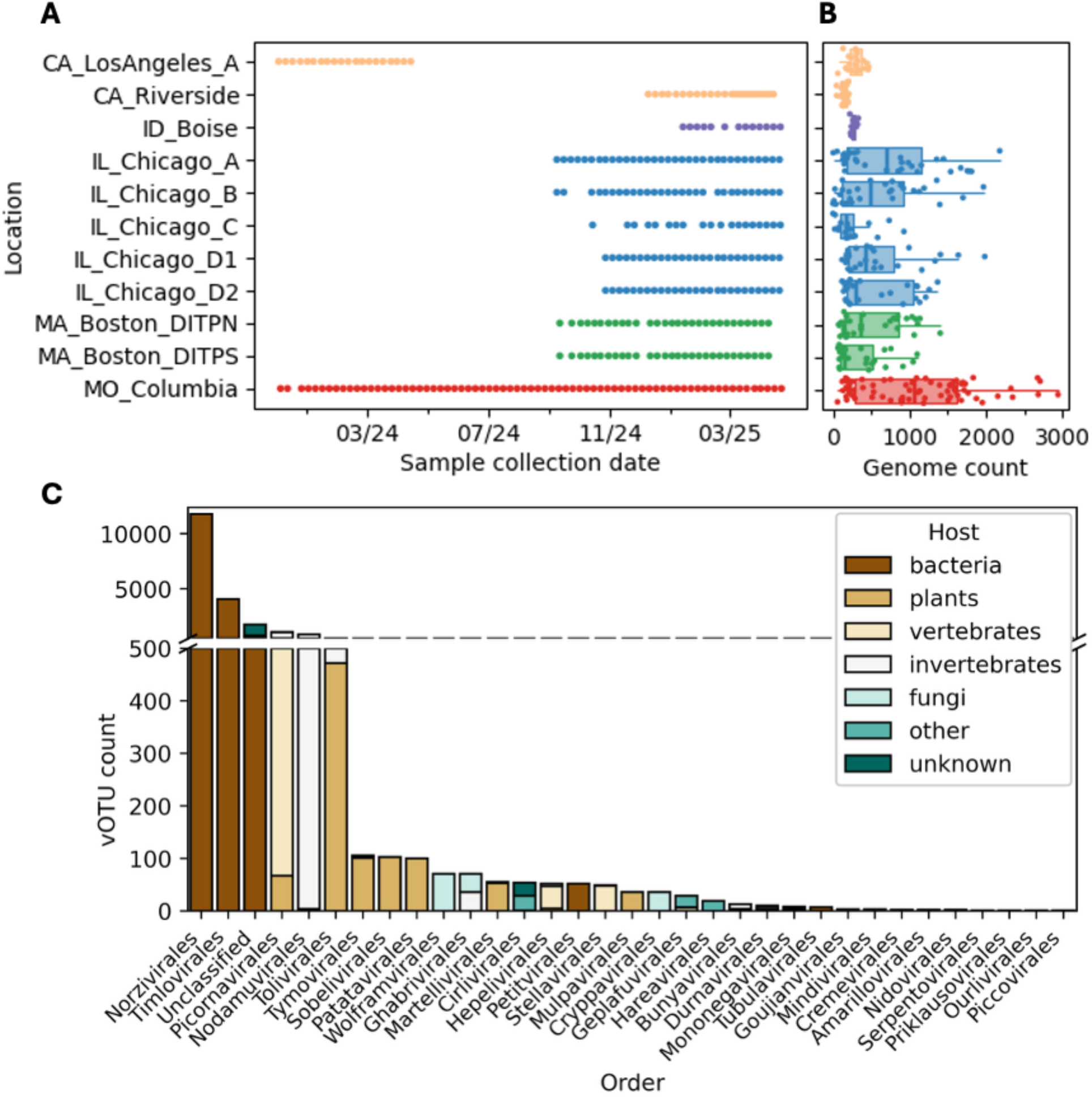
Sample, assembly, and database overview. **(A)** Metagenomes in this study were sequenced for 321 wastewater samples collected over time (x-axis) and at 11 sites in 6 US cities (y-axis). **(B)** Counts of near-complete viral genomes identified from each single metagenomic assembly across all sites. Data are colored by US states including: California (orange), Idaho (purple), Illinois (blue), Massachusetts (green), and Missouri (red). **(C)** Count of vOTUs within WVDB that belong to each viral order. Bars are colored by the predicted host for each vOTU, and “other” includes vOTUs classified to families for which the ICTV virus properties table lists multiple hosts. Hosts are shown as “unknown” where no order- or family-level taxonomic call was made and the vOTU had no genus-level BLASTN hit against NCBI core-nt.

### 2.1 Genome taxonomy, predicted host, and novelty of vOTUs in WVDB

We annotated vOTU taxonomy and predicted hosts (**Figure 1C**) using an ensemble of methods (**Figure S2C**). A total of 32 orders of viruses were recovered spanning prokaryotic and eukaryotic hosts. Of the 21,015 vOTUs, 20,671 (98%) represented RNA viruses, including 16,749 vOTUs that infect bacteria, among which 16,601 (79%) were members of the recently-defined +ssRNA bacteriophage class, Leviviricetes^26^ (most within orders Norzivirales and Timlovirales). Over eight percent could not be classified to the order level, most of them within bacteriophage-containing viral classes (**Figures 1C, S4A**). Within the vOTUs predicted to infect eukaryotes, 587 (2.8%) were predicted to be vertebrate-infecting, predominantly from families Picornaviridae (n=412), Astroviridae (n=48), Caliciviridae (n=43), and Hepeviridae (n=42). Meanwhile, 157 were predicted to infect fungi (including members of the Wolframvirales, Ghabrivirales, Cryppavirales and others), and 944 were plant infecting (including members of the Tolivirales, Sobelivirales, Tymovirales, Patatavirales, Picornavirales, Martellivirales, and others).

Further, since our ensemble methods predicted hosts only at higher taxonomic levels, we used species-level BLAST hits to NCBI core-nt to assign vOTUs as human-infecting, and recovered known enteric viruses such as *Norovirus norwalkense* (n=25), *Sapovirus sapporoense* (n=11), *Enterovirus alphacoxsackie* (n=1), *Enterovirus betacoxsackie* (n=1), *Enterovirus coxsackiepol* (n=3), *Astrovirus* spp. (n=14), and *Kobuvirus* spp. (n=3) (**Table S1**). Human respiratory viruses detected previously by read-based methods were generally too low in abundance for whole-genome assembly ^14,20^.

To assess the novelty of genomes in WVDB, we searched against other publicly available databases. Just 407 vOTUs had a BLASTN hit to NCBI core-nt at the species-level (>95% identity and >85% alignment coverage), while 1453 had a genus-level hit (70% identity and >85% alignment coverage) (**Figure S4B**). We also searched against the recently released metagenomic virus databases, metaVR ^22^ and VIRE ^23^, in addition to previously published databases on the human virome ^25,27^, but found just that 3845 vOTUs (18%) could be accounted for by genus-level hits to any database (**Table S2**). We next assessed the novelty of vOTUs based on their gene content by searching predicted proteins against profiles of virus orthologous groups (VOG HMMs, see Methods)^28^. After accounting for duplicate or overlapping hits, 12.6% of all vOTUs had no hits, and 24.8% had just one hit (**Figure S4C**).

### 2.2 Representativeness of WVDB

We next aimed to quantify how well the WVDB accounted for all viruses in wastewater in terms of detectable genomes, genes, and reads. First, we constructed a collector’s curve of genome-level vOTUs using permutations of all 321 samples. Here, we reported a genome within a sample if the genome had ≥50% coverage breadth and at least 1x coverage depth in the sample. This curve suggested that, across all samples, a plateau in new genome acquisition had been reached at around 100 samples (**Figure 2A**, black dashed line). However, we suspected that overrepresentation of samples from the Columbia site in this dataset may have artificially biased the collector’s curve. Indeed, site-wise curves using the number of new vOTUs present on actual sample collection dates revealed that the Columbia site had reached a plateau while sites with fewer sampling dates (e.g. Boise) remained in the linear phase (**Figure 2A**, colored lines). Overall, observed vOTUs were highest in the Chicago, IL city sites and lowest in Riverside, CA and the Chicago airport site (IL_Chicago_C) (the latter reached just 4924 vOTUs detected after 19 samples collected).

**Figure 2.**
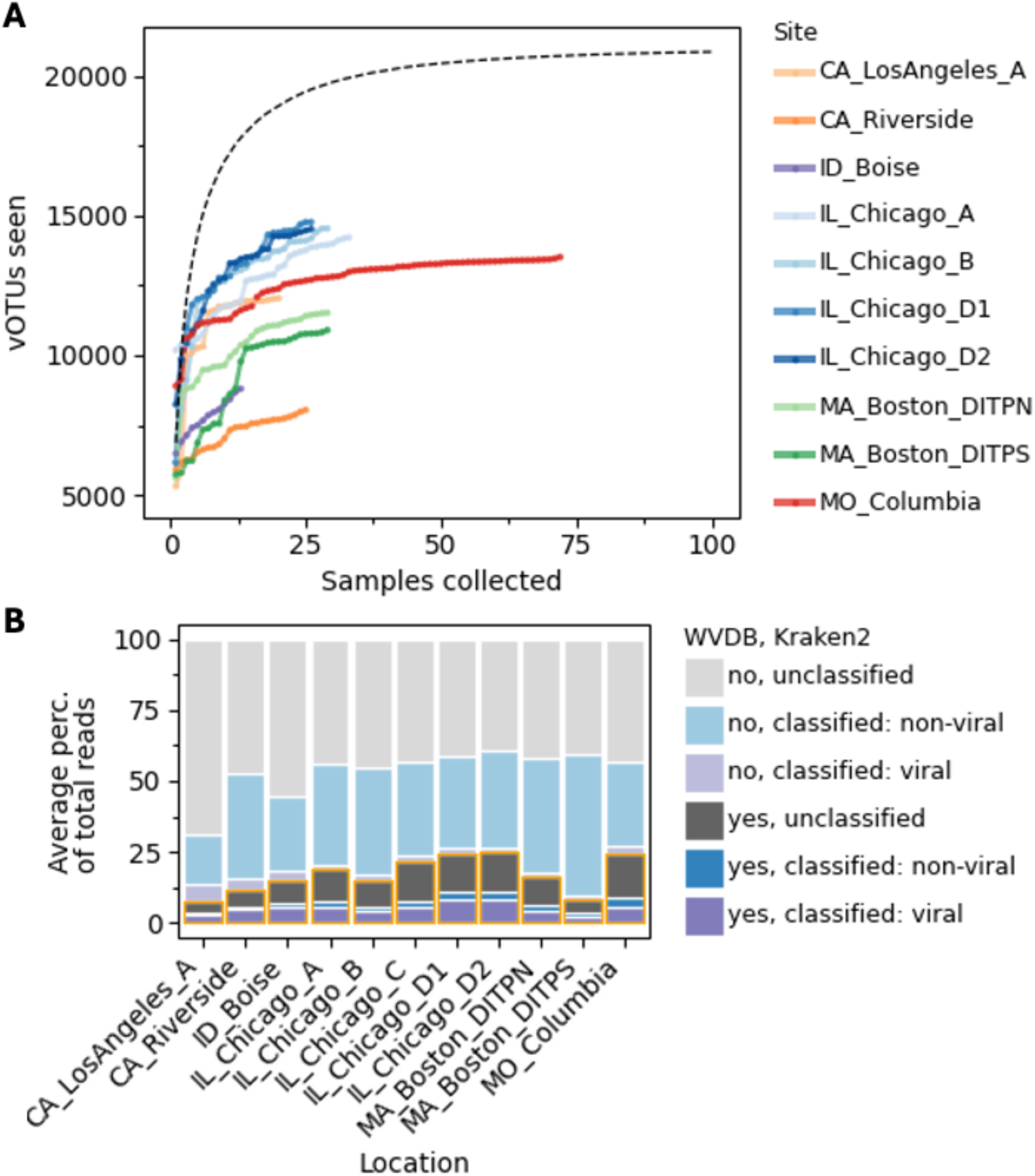
**(A)** A collector’s curve was made with 200 random permutations of the sample order across all 321 samples aggregated (black dashed line), and real-time collector’s curves show the accumulation of newly detected vOTUs over time at each site (colored lines, Chicago airport site excluded). The x-axis represents the number of samples aggregated (for the aggregated curve) or the count of samples collected at each site, ordered chronologically (for the site-specific curves). Collection of a vOTU genome from a given sample was defined as coverage was >50% breadth and >1x trimmed mean depth. **(B)** Percentages of reads classified by kraken2 as viral (purple), nonviral (blue), or unclassified (grey), and mapped (dark), or not mapped (light), to WVDB. Total average percentages of mapped reads per site are boxed in orange.

Since WVDB contains only *de novo* assembled genomes that are near-complete, it is likely missing near-complete genomes that cover the majority of a reference genome but not enough to be assembled. Thus, to record viral genomes that may not have assembled into single contigs, we compared WVDB genomes to a reference-based mapping approach (EsViritu)^27^. EsViritu identified 294 reference genomes with >90% coverage breadth (after deduplication across all samples), and produced mapping-based consensus sequences for these genomes. Of these, 129 matched to vOTUs in the WVDB (95% identity and 85% query coverage) (**Figure S5A**). The EsViritu consensus genomes not found in WVDB included 7 SARS-CoV-2 genomes, each of which contained a low-coverage region that likely broke the assembly (**Figure S5B**), resulting in exclusion from WVDB.

Likewise, to account for viruses whose genomes were incomplete but contained signature RdRp genes, we extracted assembled RdRp genes from each sample. After clustering the RdRp genes to 90% amino acid similarity over 75% of the predicted protein length representing species/vOTU level clustering, a total of 199,032 RdRp genes were recovered. At the taxonomic level, there were seven additional viral orders represented in RdRp genes from these samples that were not represented in WVDB (**Figure S3B**).

Lastly, we examined the fraction of reads per sample that mapped to WVDB and explored whether use of WVDB improved our ability to classify wastewater sequencing reads overall. Read mapping of individual samples to WVDB showed that the database accounted for an average of 18.3% (range 0.4 - 57.2%) of all clean, non-ribosomal RNA reads per sample (**Figure 2B**, orange outline, and **Figure S6**). The wide range is likely due to sample-to-sample variability in viral content and uneven representation of total viromes from each site and city in WVDB (**Figure 1B**). Further, we compared classifications by Kraken2 with the NCBI core-nt database to read-mapping against the WVDB on a per-read level (**Figure 2A**, bar fill color). On average among all samples, 7.5% of reads were classified as viral by Kraken2 (range 0.4 - 44.7), and WVDB typically captured most of these reads (mean 5.3%, range 0.0 - 30.3%). Notably, on average 56% (range 23.1 - 83.6%) of total reads were unclassified by Kraken2 (**Figure 2B**, light and dark gray bars), but within this set, 10.9% (range 0.0 - 40.3) mapped to WVDB (**Figure 2B**, dark gray bars), representing a direct improvement in read classification when using WVDB.

### 2.3 A genome-informed view of the wastewater virome composition

An initial analysis of the viral-classified reads with Kraken2 (**Figure S7**) showed that an average of 27.1% could not be classified to the order level, while the most abundant order at most sites was Martellivirales, 27% (which contains the plant-infecting family Virgaviridae). Given the improvement in proportion of reads accounted for with WVDB, we re-characterized the predicted host and taxonomic composition of each site based on classifications of vOTUs in WVDB (**Figure 3A and 3B**). Focusing here on only the reads that mapped to WVDB, a smaller fraction mapped to vOTUs classified within Martellivirales (12.4%), just 7.1% were unclassified at the order level, and the ssRNA bacteriophage order Norzivirales averaged 45.5%. Overall, bacteriophage were the most abundant vOTUs across all sites, followed by those predicted to infect plants, invertebrates, and vertebrates.

**Figure 3.**
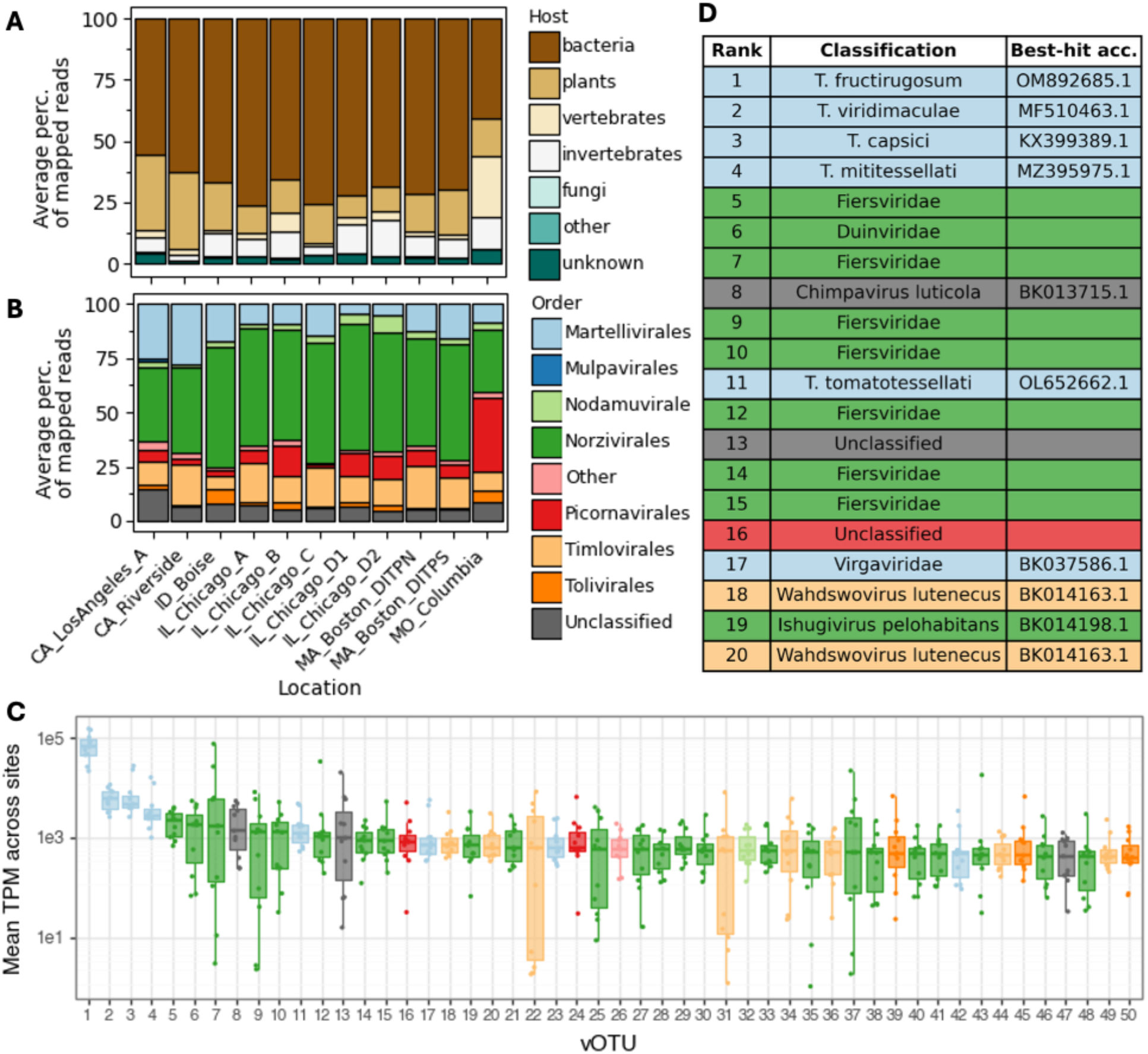
Out of WVDB-mapped reads, percentages of reads mapped to vOTU genomes within each host category **(A)** and viral order **(B)**. Percentages were averaged among samples within each site (bar). **(C)** Abundances (y-axis) of the top 50 vOTUs (x-axis) present at least once across all sites. TPM (relative abundance) was averaged over time for each vOTU within each of the 11 sites, and boxes indicate the median and interquartile range of mean TPM across sites. vOTUs are rank-sorted in descending order of median(mean TPM). Boxes are colored according to the order-level classification of the vOTU, as in (B). (**D**) For the top 20 vOTUs shown in (C), the best genus- or species-level BLASTN hit to NCBI core-nt and associated accession (if available) is shown, colored by order-level classification, as in (B).

To identify the most abundant and ubiquitous WVDB viruses in all samples, we then focused on vOTUs present at least once in all sites. The mean transcripts per million (TPM) for each of these vOTUs was calculated for each site (**Figure 3C**), and rank sorting revealed that the four most abundant vOTUs corresponded to known *Tobamovirus* species (plant-infecting), consistent with previous studies ^7^. Beyond the *Tobamovirus* spp., some vOTUs displayed large site-to-site variability in abundance, especially some of those classified within the orders Norzivirales and Timlovirales.

### 2.4 Genome-enabled spatial dynamics of the wastewater virome

When using WVDB to examine the wastewater virome across space, clear site-specific patterns were observed, as well as strong similarities within cities or regions (**Figures 4A and S8**). Notably, the Chicago airport site (IL_Chicago_C) clustered centrally amongst the city sites, perhaps due to its lower vOTU richness (see Section 2.2). The importance of site was much greater than that of sampling date although both were significant (PERMANOVA R^2^(site)=0.589, R^2^(date)=0.026, R^2^(site-date)=0.102, p<0.001 for all; note that sample counts were uneven across sites). This suggests a relatively consistent virome profile within each site. The majority of vOTUs occurred in multiple, but not all, sites. The number of vOTUs that occurred in all eleven sites was similar to the number of vOTUs observed in only one site (**Figure 4B**).

**Figure 4.**
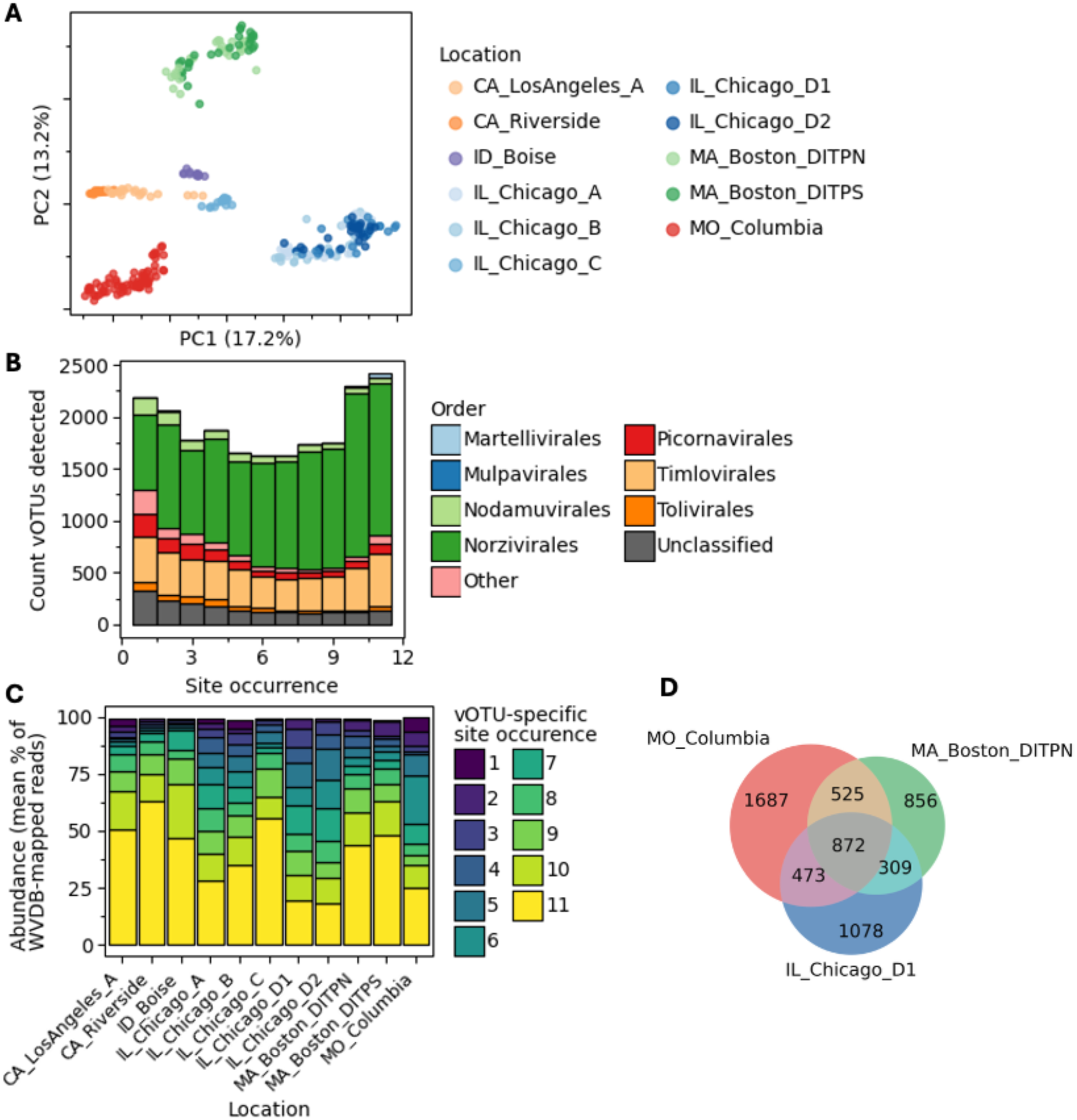
**(A)** PCoA plot using CLR-transformed relative abundance (TPM) to calculate Aitchison distance. Samples are colored by site and state. See Figure S4 for additional ordinations. **(B)** vOTUs were categorized by the number of sites at which they were present on ≥1 date (x-axis) and the number of vOTUs in each category is shown on the y-axis. Presence of each vOTU in each sample was defined as >50% coverage breadth and 1x coverage depth on at least one date per site). vOTUs are colored by virus order-level classification. **(C)** For each site (x-axis), the average breakdown of WVDB-mapped reads according to whether they map to core-like vOTUs or unique vOTUs (bar segments). vOTUs are categorized as in (B) according to site-wise presence (1 to 11 sites), and bar segments are colored according to category. **(D)** Overlaps between the recurrent viromes for each of three well-sampled sites in the dataset. Here, “recurrent” included vOTUs present on ≥90% of all sampling dates at a given site for all sites within the city.

To examine how much of the virome was accounted for by site-specific versus widespread vOTUs, we categorized vOTUs according to the number of sites they were present at and then broke down the total mapped reads by which category of vOTU they mapped to (**Figure 4C**). In all sites, the ubiquitous virome was more abundant than the site-unique virome. Specifically, in the California, Boise, and Chicago airport sites, the widespread virome (10-11 sites) accounted for over half of the total mapped reads, on average. Meanwhile, Chicago and Columbia had higher average abundances of vOTUs present in 6 or fewer sites. This result may be partly a reflection of the better representation of the database for Columbia and Chicago relative to the other sites (**Figure 1A & B**).

We next examined whether certain occurrence patterns differed by taxonomy within selected viral orders. Martellivirales vOTUs were predominantly classified within the plant-infecting Virgaviridae family and appeared to be highly ubiquitous - the majority of these vOTUs occurred in all 11 sites (**Figure S9A**). Meanwhile, Nodavirales vOTUs were predominantly unclassified at the family level and more were site-specific than were ubiquitous (**Figure S9B**). The Picornavirales showed multiple trends. First, vOTUs within the Calciviridae, which contain *Norovirus spp.* and *Sapovirus spp.* were largely widespread. Meanwhile vOTUs within the Picornaviridae and unclassified Picornavirales ranged from widespread to site-specific (**Figure S9C**).

Because viromes appeared to cluster by location (**Figure 4A**), we compared the recurrent viromes within specific sites in Columbia, Boston, and Chicago using a threshold of ≥90% occurrence frequency across sample dates within a site (**Figure 4D**). Out of a total of 5800 vOTUs that were determined to be recurrent in any of the three sites, 872 (15%) were recurrent in all three, while 2,179 (38%) were recurrent in at least two sites.

## 3. Discussion

Our findings are well-aligned with previous wastewater RNA sequencing efforts showing bacteriophage make up the majority of RNA viruses, while plant viruses are highly abundant. We demonstrate that reference-based classification with existing databases (NCBI core-nt) is insufficient to capture this diversity. We further show that with the vOTU genomes in WVDB, we are able to classify considerably more viral reads than with existing databases. This classification enables us to distinguish samples by site and generate genome-resolved core virome information.

### 3.1 Potential uses and benefits of the database

Applications of the database and associated abundance information range from fundamental to applied. The genomes in the database can be used to fill in viral phylogeny, particularly for RNA phages and novel vertebrate and invertebrate-infecting viruses. Relatedly, these genomes can be used to expand viral protein space and build new profiles (HMMs or other) like those in VOGDB^28^ for improved identification and classification of these groups. Given that wastewater represents a mixed-species and, more importantly a mixed-strain sample, genome resolution can be used to track viral population heterogeneity in wastewater, within individual vOTUs. For example, SNV frequencies and rates of change over time could be measured for genomes of interest, expanding on work done with SARS-CoV-2 and other human-infecting viruses. Practical applications of the database include use as a “background” against which to check new quantitative PCR assays for potential false positives prior to use on wastewater samples. Additionally, this background could be used to build probe panels for *in vitro* depletion of the abundant baseline wastewater virome prior to sequencing.

Most critically, WVDB represents a wastewater virome baseline that covers a larger fraction of the read data than existing reference databases. Use of WVDB to complement these references may enable the detection of novel viral sequences by removing previously unclassified reads from consideration as novel threats. Incorporation of WVDB into ongoing real-time analyses of ultra-deep sequencing may speed up this detection.

### 3.2 Challenges and limitations

Although WVDB provides an expanded view of viruses in wastewater, it remains a partial view. While vOTU collector’s curves appear to plateau once a given site has been deeply sampled (Section 2.2), the rigorous nature of the vOTU reconstruction and selection process employed here artificially limits the ceiling on what can be collected. Viral genomes that will be missed by our workflow include viral “dark matter”, such as sequences that are not flagged as viral by geNomad and/or not estimated to be near-complete by checkV. Additionally, the single-contig approach employed here will not capture complete genomes for segmented viruses, instead keeping only the segment containing key marker genes for completeness such as the RdRp gene (e.g. the large segment for Bunyavirales). Lastly, our approach also does not include singletons that represent rarer viruses as our priority was recovering high quality genomes: due to the rigorous chimera trimming protocol, each vOTU must assemble completely at least twice for inclusion in WVDB.

An obvious further limitation is that WVDB will not include vOTUs whose abundance in wastewater is always below what is needed to achieve 1-2x coverage across the entire genome. These vOTUs will not assemble into a single contig in single-sample assemblies, necessitating a different approach. Established methods such as probe-capture enrichment ^9,16,27,29,30^ or DNA-only sequencing could be used to collect specific genomes of interest. It has also been shown that different methods of wastewater sample processing^30,31^ can affect viral abundances ^30,31^, and these biases could perhaps be used to deliberately collect a greater diversity of viruses from biological replicate samples. Lastly, co-assembly of multiple samples (perhaps only reads that don’t map to WVDB) could be used to recover additional viral genomes. While this risks the creation of chimeras, it may enable the assembly of low-abundance viruses.

Another challenge is in the area of virus host assignment. Although tools have been developed to link bacteriophage to bacterial hosts in metagenomic data ^32^, host assignment for non-bacterial viruses remains complex. Machine-learning based methods such as RNAVirHost are limited by the quality and extent of the training data available for model construction ^33^. There remains a high degree of underexplored viral diversity, particularly for microbial eukaryotes, and non-vertebrate eukaryotes in general (including contributors to human diets). Another complication is that because of the exploratory nature of host virome studies, viral genomes recovered from those works cannot be conclusively tied to the host rather than to the host’s diet or microbiome. This leads to GenBank entries where the “host” field may be incorrect. Especially notable are plant-infecting viruses that are found in vertebrate fecal metagenomes. Our ensemble approach to host prediction identified disagreements and gaps between methods, and host predictions provided for WVDB members should be interpreted with caution. Possible routes for improvement of host assignments include 1) sequencing of microbial eukaryotes from pure culture, co-culture, or single-cell sequencing, 2) cross-linking host and viral DNA prior to sequencing ^34^ (this technology does not currently enable RNA sequencing), 3) bioinformatic linking via time series abundances of host and virus genomes captured through parallel sequencing campaigns (e.g. eukaryotic 12S, 18S or ITS sequencing). Overall, we note that wastewater-derived vOTUs cannot be definitively linked to hosts without additional evidence.

### 3.3 Future updates and expansion

As the CASPER data collection effort is ongoing, WVDB will continue to iteratively expand. The initial version of WVDB was built with data collected through May 2025, and in the intervening time, CASPER has expanded to >40 sites and >2000 samples. We anticipate that including vOTUs generated over this longer time series and across more sites will result in a more comprehensive database. Columbia served as the pilot site for this work and therefore has many more samples included in WVDB than other sites. Partly for this reason, a higher fraction of reads from Columbia samples are accounted for by WVDB. We expect that the metric of viral reads accounted for in each sample will increase as more data is included in WVDB. Additionally, the extended time series dataset will also improve our ability to perform temporal analyses for non-human viruses of interest. Lastly, the workflows built here can be used to produce vOTUs for inclusion in future versions of WVDB, with the goal of automation across incoming sequence data. This may also trigger alerts for detection of novel viruses in real time.

## 4. Methods

### 4.1 Data collection

All raw read data used in this study are publicly available at NCBI SRA, BioProject numbers PRJNA1247874 and PRJNA1198001. Sample collection and processing was previously described in detail by Justen *et al.*^14^ Rushford *et al.*^20^, and Grimm *et al.* ^21^ and all samples except CA_LosAngeles_A in this study were processed at the University of Missouri. Site names shown here are matched to those manuscripts but include the state and city name where possible (e.g. “CHI-D1” is “IL_Chicago_D1” in this manuscript).

Briefly, all wastewater was collected as 24-hour composite samples except at IL_Chicago_C, where Moore swabs were collected from a manhole serving an airport. After solids removal, viruses were concentrated using a polyethylene glycol (PEG) precipitation method followed by extraction with the QIAamp Viral RNA Mini Kit (Qiagen, 52906). Carrier RNA and DNase treatment were omitted. Some samples from Columbia, MO, in early 2024 underwent parallel concentrations and extractions with additions to the protocol. Here, we used only the subset of replicates whose protocol had no modifications, except for samples collected between January 16 and February 20, 2024, which were treated with 10,000 ng RNase A and 30 mM EDTA before filtration (RNase A was inactivated prior to extraction). Sequencing libraries were prepared with the Illumina Stranded mRNA Prep, omitting mRNA capture. Where there were multiple libraries or sequencing runs for the same sample, this study used only the libraries or runs sequenced to the depth of ∼1 billion reads with read length 150 bp or 100 bp (as available). Samples from CA_LosAngeles_A were filtered for solids removal, concentrated with 10-kDA Amicon filters (Millipore-Sigma), then RNA was extracted with an Invitrogen PureLink RNA kit with DNase. CA_LosAngeles_A libraries were prepared with an Illumina RNA Prep with Enrichment Kit and sequenced at a depth of ∼2 billion paired-end 150bp reads.

### 4.2 Read processing

Reads were quality trimmed and adapters were removed with fastp (v0.24.0) ^35^, using settings to filter reads below 50 nt in length and trim homopolymers and low-complexity sequences. (-l 50 --dont_eval_duplication --trim_poly_g --poly_g_min_len=5 --poly_x_min_len=10 --low_complexity_filter 30 -y). Next, ribosomal RNA was removed using bbduk (v39.13) ^36^ with default parameters against a custom rRNA database (for more information, see https://www.biostars.org/p/159959/#159960). Finally, human reads were removed using sra-human-scrubber (v2.2.1) ^37^ with default parameters. Read statistics were collected using seqkit (v2.9.0) ^38^. Reads were classified with Kraken2 (v2.17.1)^39^ using the publicly available NCBI core-nt database (version 10/15/2025). Kraken2 outputs were aggregated using KrakenTools (v1.2.1)^40^ scripts kreport2mpa.py to summarize read counts and combine_mpa.py to combine reports across samples.

### 4.3 Assembly, dereplication, and curation of the set of viral genomes

Processed reads were assembled with MEGAHIT (v1.2.9) ^41^ to produce a single assembly for each sample. Viruses were identified with geNomad (v1.11.0, database v1.9) ^42^ and genome quality was assessed using checkV (v1.0.3, database v1.5) ^43^ in end-to-end mode. High-quality viral genomes were defined as those on contigs > 2 kbp identified by geNomad as viral with checkV completeness > 90%. Proviruses were trimmed by checkV. The high-quality viruses and trimmed proviruses were collected from single assemblies and clustered to the vOTU level using vclust (v1.3.1) ^44^ with the Leiden algorithm and thresholds of 95% average nucleotide identity (ANI) and 85% query coverage.

A custom workflow was developed to detect and remove potential chimeric sequences in vOTUs caused by misassemblies (**Figure S2**). First, the two longest genomes in each cluster were identified, pairwise aligned with NUCmer (v4.0.1) ^45^ followed by show-coords (options -r -c -l -L 1000 -I 90 -T). If this resulted in more than one alignment per pair, the longer alignment was chosen. The coordinates of the alignment were used to trim the longest genome (the cluster representative), to remove potential chimeric regions from the 5’ and 3’ ends. The trimmed genomes were reassessed for completeness with checkV. If the trimmed genomes were ≤90% complete, a second iteration of alignment and trimming was conducted using the second- and third-longest genomes within each vOTU cluster (where available). Next, three categories of vOTUs were passed into a parallel workflow: singletons, those incomplete after the first round of trimming whose vOTU cluster contained <3 members, and those incomplete after the second round of trimming. These vOTU representatives (longest genomes) were compared with BLASTn (2.16.0+, megablast default, flags -max_target_seqs 10 -perc_identity 90) to RefSeq Virus (v232) plus the curated EsViritu reference database of viral genomes (v3.2.4) ^46^ and to metaVR (IMGVR-5) ^47,27,22^. Each BLAST output was parsed using blastani.py (https://github.com/snayfach/MGV/blob/07c88e92f14ffdf61ce925a0450c952c34229f5a/ani_cl uster/blastani.py), and filtered to reference sequences with ≥85% query coverage or ≥85% target coverage. Of these, the reference sequence with the highest query coverage was selected for alignment to the vOTU representative. Alignment and trimming were conducted as described above, and completeness was re-evaluated with checkV. If a vOTU was complete after trimming against references from multiple databases, the version trimmed against the RefSeq Virus plus EsViritu database was preserved.

Finally, genomes >90% complete after trimming were clustered again with vclust as above to collapse redundancy. Quality checks with checkV and geNomad identified a single vOTU with a whole-genome inverted duplication (kmer-freq=2) and another genome that geNomad identified as a provirus. These genomes were manually trimmed and added back to the final set.

### 4.4 vOTU taxonomic annotation and host assignment

Annotation of the final viral genomes was conducted with an ensemble of tools including checkV, geNomad, RdRpCATCH ^48^, RNAVirHost ^33^, and BLASTn ^47^ against a decontaminated version of NCBI core-nt^49^ and metaVR (IMGVR-5) ^22^. Species-level and genus-level BLAST hits were defined as >95% ANI and 70% ANI, respectively, across 85% of the query ^50^. BLASTn searches were conducted using parameters -evalue 1e-3 -max_target_seqs 10, followed by parsing with blastani.py (https://github.com/snayfach/MGV/blob/master/ani_cluster/blastani.py) at the above thresholds. The best BLAST hit per vOTU was chosen as the hit with the highest product of query coverage and percent identity.

To determine host assignment, three parallel methods were executed and then combined according to a decision tree model (**Figure S2C**) designed to minimize error and the need for manual review. In preparation, the taxonomic order and family of each vOTU was extracted from the RdRpCATCH annotation. Where order or family was unclassified, the taxonomy from geNomad was used instead. Order-level information was used as input for the RNAVirHost classifier with default parameters. Family level information was used to perform a look-up against the ICTV Virus Properties table^51^ which lists host(s) for reference viruses belonging to each family. Lastly, if the vOTU had a genus- or species-level BLASTN hit to NCBI core-nt, NCBI Entrez was used to extract “host” and “isolation source” for the hit accession from GenBank. If RNAVirHost and ICTV methods agreed, then the RNAVirHost was assigned. If they disagreed or couldn’t assign a host, the BLAST hit method was used as a tiebreaker. Overall, the tree preferred whichever method(s) produced an assigned host over an unassigned host and preferred the ICTV method over RNAVirHost. Finally, if family- and order-level information was missing for a vOTU, but the class was Leviviricetes, Caudoviricetes, Vidaverviricetes, or Faserviricetes, the host assignment was updated to “bacteria”.

Analyses with VOGDB began with proteins predicted with Prodigal-gv (V2.10.0-gv). Next, hmmscan from the HMMER suite (3.4) was used to search to proteins and the domtlbout format was parsed in python in the following way: 1) hits were prefiltered at e-value ≤ 1e-3; 2) same-query same-HMM hits were merged, allowing no more than 20 amino acids between hit regions and keeping score information from the best-hitting chunk; 3) hits were strictly filtered at e-value ≤ 1e-5 and target HMM coverage ≥ 70%; 4) Overlapping hits to different HMMs (if ≥50% coverage overlap) were collapsed, keeping the best hit per region.

### 4.5 Relative abundance analyses

To determine abundance of each vOTU in each sample, reads were mapped against the WVDB using bowtie2 (v2.5.4, with parameters --no-unal --no-discordant --no-mixed --sensitive) ^52^ and output was converted to sorted bam files with samtools (v1.21). Coverage calculations were performed with coverM (v0.7.0) to return read count, covered bases, contig length, trimmed-mean coverage depth and transcripts per million (TPM). CoverM results were prefiltered to remove vOTU-by-sample combinations with fewer than 3 reads mapped to diminish the effect of stochastic or nonspecific mapping results.

### 4.6 RdRp analyses

All contigs greater than 500 bp were annotated as described above. Gene prediction was performed using Prodigal-gv, and the resulting protein sequences were searched against a collection of RdRp HMM databases implemented in the RolyPoly pipeline, including geNomad, RdRp-Scan, Pfam_A_37, TSA_Olendraite, RVMT, and NeoRdRp v2.1 ^53,42,54–58^. Only top-scoring hits were retained for downstream analysis. Candidate genes were subsequently filtered based on the presence of the conserved RdRp A, B, and C domains, as defined by the PALM motif database within PalmDB, since these motifs represent the core catalytic features of viral polymerases. We further required that these domains occurred in either A–B–C or C–A–B order, reflecting known canonical and permuted arrangements observed in viral RdRps. Sequences containing partial domains or alternative domain arrangements were discarded. Finally, the retained sequences were clustered at 90% amino acid identity over 75% alignment coverage using MMseqs2 (version 18d8ddc2f468b8a00cec9addb3d2328eb790ac9e)^59^.

### 4.7 Data analysis

Data was organized and analyzed using Python3 with pandas (v2.2.3) ^60^ and visualized with libraries plotnine (v0.13.6) ^61^ and matplotlib (v3.9.2) ^62^. Where relevant, vOTU presence in a sample was defined as ≥50% coverage breadth and ≥1x trimmed-mean coverage depth based on mapped reads from that sample. Site-wise presence required presence in at least one sample from a given site. Recurrent viromes were defined as vOTUs present in ≥90% of all samples from a given site.

Beta-diversity analyses were performed by CLR-transforming TPM values and calculating Aitchison distances for PCoA plots and PERMANOVA testing. PERMANOVA was performed with 999 permutations using the rpy2 library in Python to access the vegan *adonis* function from R. Variables were listed in the order: site, date, site-date interaction. Similar results were produced using CLR-transformed raw read counts instead of TPM (**Figure S8**).

### 4.8 Data and code availability

Data are available at https://zenodo.org/records/20276352. Viral database building workflows are available at: https://github.com/IL-wastewater/WVDB_workflows.

## 5. Acknowledgements

We thank Livermore Computing (LLNL), the University of Missouri Genomics Technology Core (MU), and operators and staff at wastewater treatment facilities including the Riverside Water Quality Control Plant and the MWRA Central Laboratory Violet Team. Chicago samples were provided by the Metropolitan Water Reclamation District of Greater Chicago and the Chicago Department of Aviation in collaboration with the Chicago and Illinois Wastewater Surveillance Systems. Part of this work used the San Diego Supercomputer Center’s Expanse, through allocation BIO240238 to J.A.R. from the Advanced Cyberinfrastructure Coordination Ecosystem: Services & Support (ACCESS) program, which is supported by U.S. National Science Foundation grants #2138259, #2138286, #2138307, #2137603, and #2138296. Funding for this work was provided by Inkfish and SecureBio; funding to RSK, MS, NR, AE, JEA and PSGC was provided by interlaboratory LDRD award #24-ERD-056/20240674ER. This work was performed under the auspices of the U.S. Department of Energy by Lawrence Livermore National Laboratory under Contract DE-AC52-07NA27344.

## 6. Author contributions

**Conceptualization:** Marc Johnson, Rose Kantor, Migun Shakya, Jason Rothman, Dave O’Connor

**Methodology:** Rose Kantor, Migun Shakya, Marc Johnson, Devon Gregory, Clayton Rushford

**Software:** Rose Kantor, Migun Shakya, Nelson Ruth, Aidan Epstein

**Validation:** Rose Kantor

**Formal analysis:** Rose Kantor, Migun Shakya

**Investigation:** Rose Kantor, Migun Shakya, Marc Johnson

**Resources:** Patrick Chain, Jonathan Allen, Marc Johnson, Dave O’Connor, Jeff Kaufman, Jason Rothman

**Data Curation:** Rose Kantor, Nelson Ruth, Clayton Rushford

**Writing - Original Draft:** Rose Kantor, Migun Shakya

**Writing - Review and Editing:** Rose Kantor, Migun Shakya, Marc Johnson, Jason Rothman, Devon Gregory

**Visualization:** Rose Kantor, Migun Shakya

**Supervision:** Patrick Chain, Jonathan Allen, Marc Johnson

**Project administration:** Marc Johnson, Rose Kantor, Migun Shakya

**Funding acquisition:** Patrick Chain, Migun Shakya, Jonathan Allen, Marc Johnson, Dave O’Connor

## 8. Statement on AI usage

AI tools were used to assist in editing of manually coded workflows and analysis scripts, directed coding, and to identify grammatical errors and inconsistencies in the manuscript. All code and text was manually reviewed and edited by the authors. No figures or text were generated by AI.

