## Supplementary Material for "A genome-resolved view of the wastewater RNA virome"

[*]These authors contributed equally

**Supplementary Material**

Tables: 2
Figures: 9

### Tables

**Table S1.** Count of vOTUs for which the best BLASTN hit to the NCBI core-nt database was reported to have host as Homo sapiens.

| BLAST hit taxonomy | vOTU count |
|---|---|
| Astroviridae | 4 |
| Mamastrovirus | 2 |
| Mamastrovirus hominis | 7 |
| HMO Astrovirus A | 1 |
| Enterovirus alphacoxsackie | 1 |
| Enterovirus betacoxsackie | 1 |
| Enterovirus coxsackiepol | 3 |
| Kobuvirus | 1 |
| Kobuvirus aichi | 2 |
| Norovirus norwalkense | 25 |
| Sapovirus sapporoense | 11 |
| Picornavirales | 3 |

**Table S2.** BLASTN hits of WVDB genomes to other public databases.

| Database* | Species-level hits (95% / 85%) | Genus-level hits (70% / 85%) | Sources | Reference | Total genomes in db | Version date |
|---|---|---|---|---|---|---|
| NCBI core-nt | 444 | 1453 | core-nt | https://ncbiinsights.ncbi.nlm.nih.gov/2024/07/18/new-blast-core-nucleotide-database/ Martí et al. 2025 (curation methods) | NA | Jul 5, 2025 |
| metaVR (IMGVR5) | 1142 | 3524 | metagenomes, metatranscriptomes, isolates, SAGs, MAGs | Fiamenghi et al. 2025 | 24,435,662 | Dec 2, 2025 |
| VIRE | 55 | 171 | SRA/ENA metagenomes | Nishijima et al. 2025 | 1,784,510 | Dec 9, 2025 |
| CHVD | 12 | 39 | Human microbiome | Tisza et al. 2021 | 45,033 | Feb 3, 2021 |
| UHGV | 18 | 71 | Human gut virome ("HQ plus") | Camargo et al. 2025 (preprint) | 873,994 | Nov 19, 2025 |

*Note that UHGV contains CHVD, and metaVR contains UHGV

### Figures

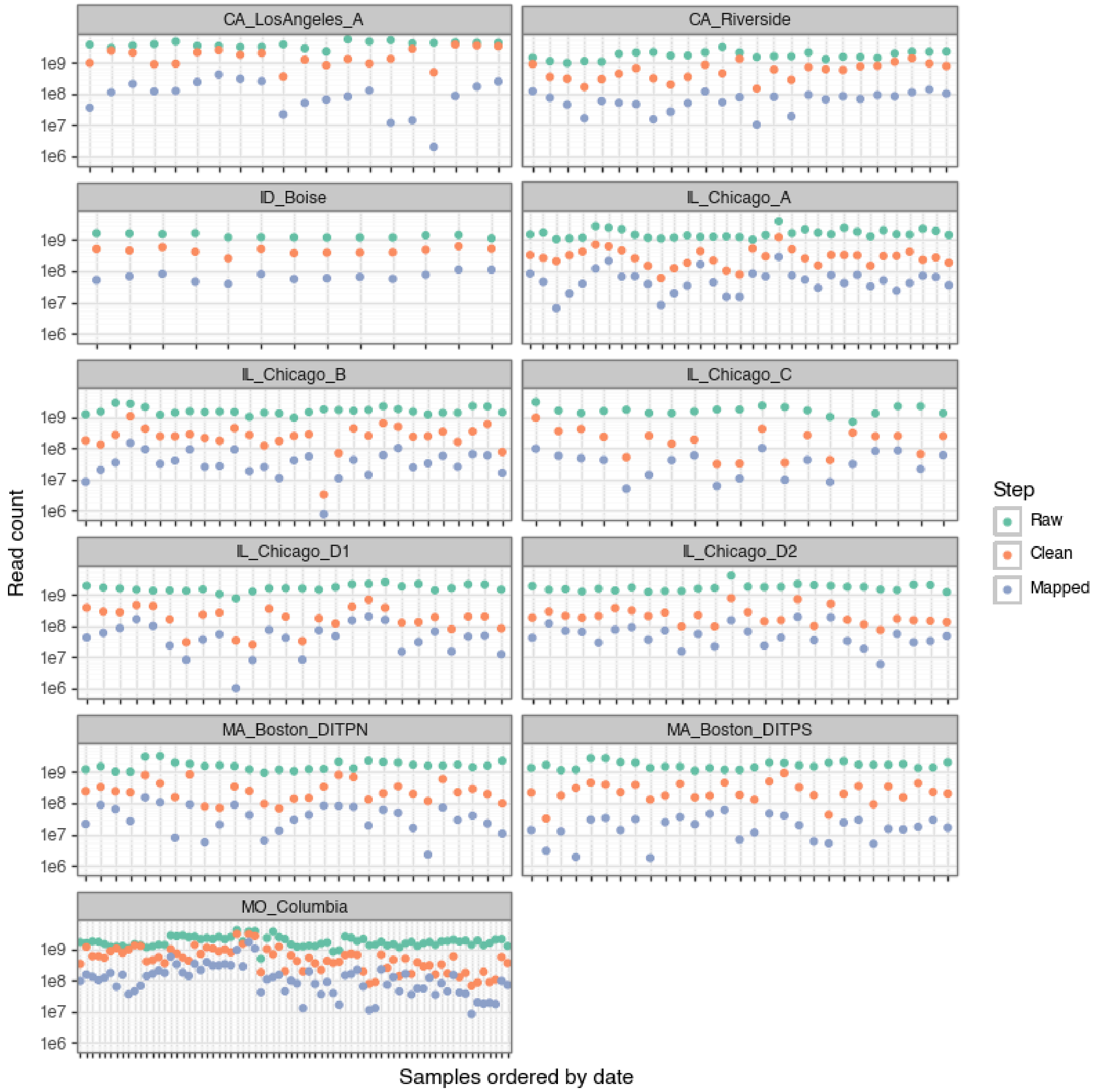


**Figure S1.** Read counts for each sample, within each site. Points are colored by raw reads (blue), processed reads produced after rRNA and human sequence removal (green), and reads mapped to the viral genome database (red). Note that samples are arranged chronologically for display purposes, but dates do not align across sites (see **Figure 1**).

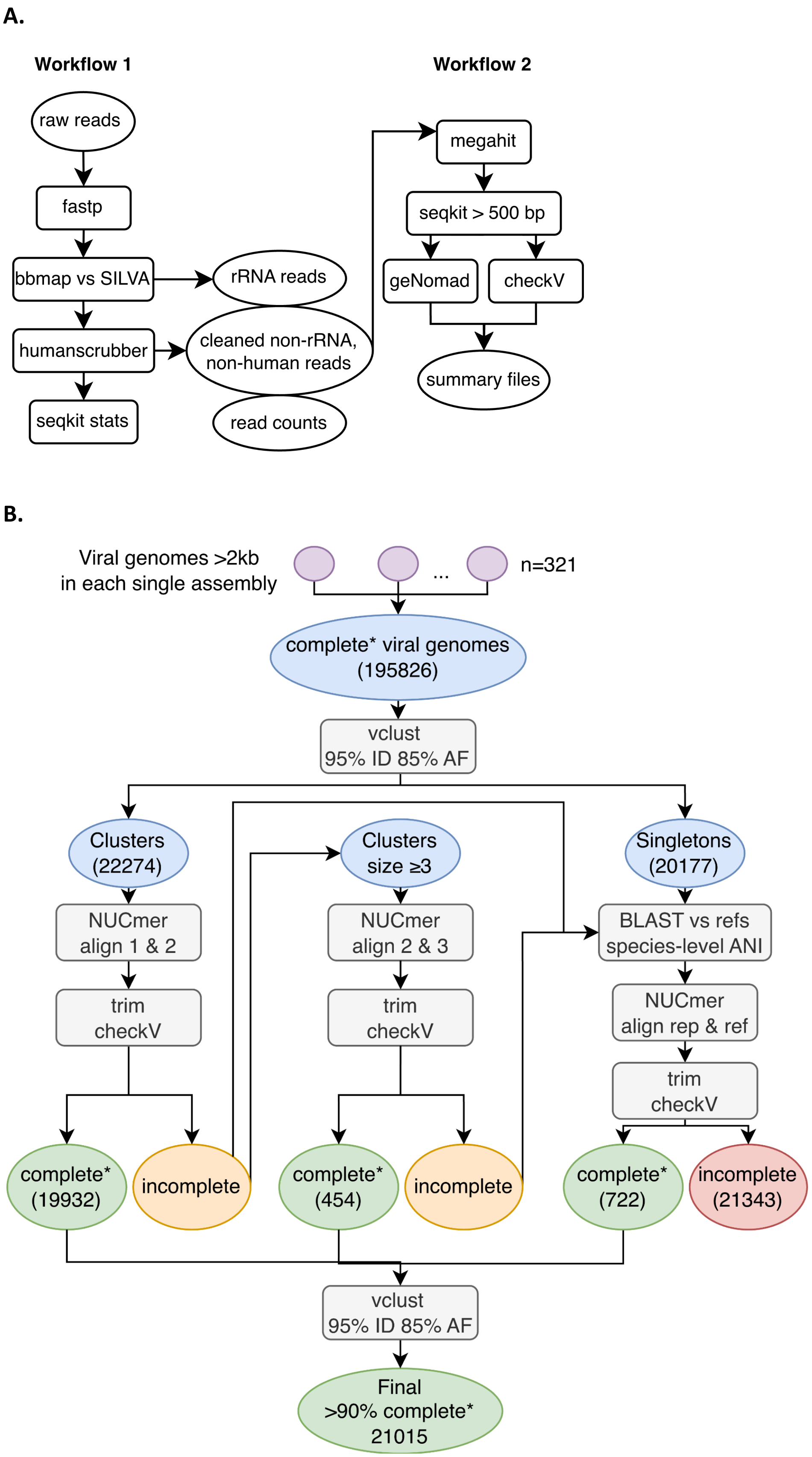
A.
Workflow 1
raw reads
fastp
bbmap vs SILVA
rRNA reads
humanscrubber
cleaned non-rRNA, non-human reads
seqkit stats
read counts
Workflow 2
megahit
seqkit > 500 bp
geNomad
checkV
summary files
B.
Viral genomes >2kb in each single assembly
...
n=321
complete* viral genomes (195826)
vclust 95% ID 85% AF
Clusters (22274)
NUCmer align 1 & 2
trim checkV
complete* (19932)
incomplete
Clusters size ≥3
NUCmer align 2 & 3
trim checkV
complete* (454)
incomplete
Singletons (20177)
BLAST vs refs species-level ANI
NUCmer align rep & ref
trim checkV
complete* (722)
incomplete (21343)
vclust 95% ID 85% AF
Final >90% complete* 21015

**C.**

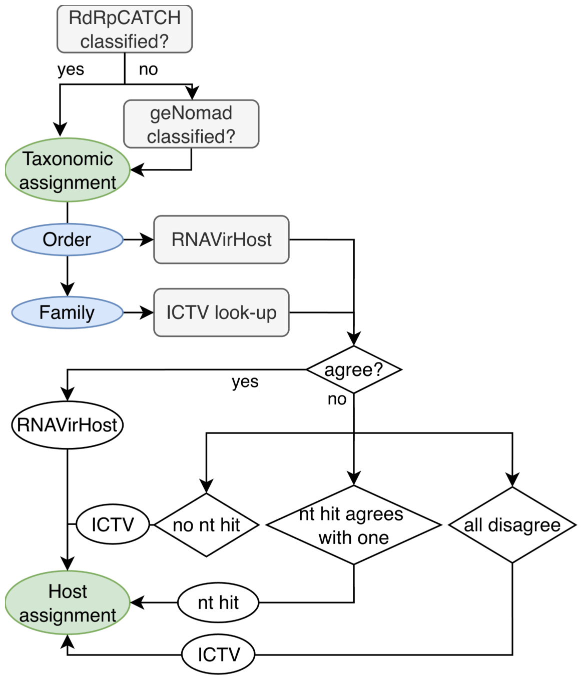


**Figure S2.** Workflows for data processing. **(A)** Nextflow workflows for cleaning raw reads and performing *de novo* assembly and viral genome detection. **(B)** Workflow for collecting and curating genomes for vOTUs in WVDB. *Genome completeness is defined as >90% based on to checkV quality summaries. See Methods for detailed description of all workflows. **(C)** Ensemble model for determination of host assignment using RdRpCATCH and geNomad for taxonomic classification, followed by RNAVirHost, look-up to the ICTV Virus Properties table ("ICTV"), and extraction of host taxonomy from NCBI GenBank entry of best BLASTN hit against NCBI core-nt ("nt hit").

**A.**

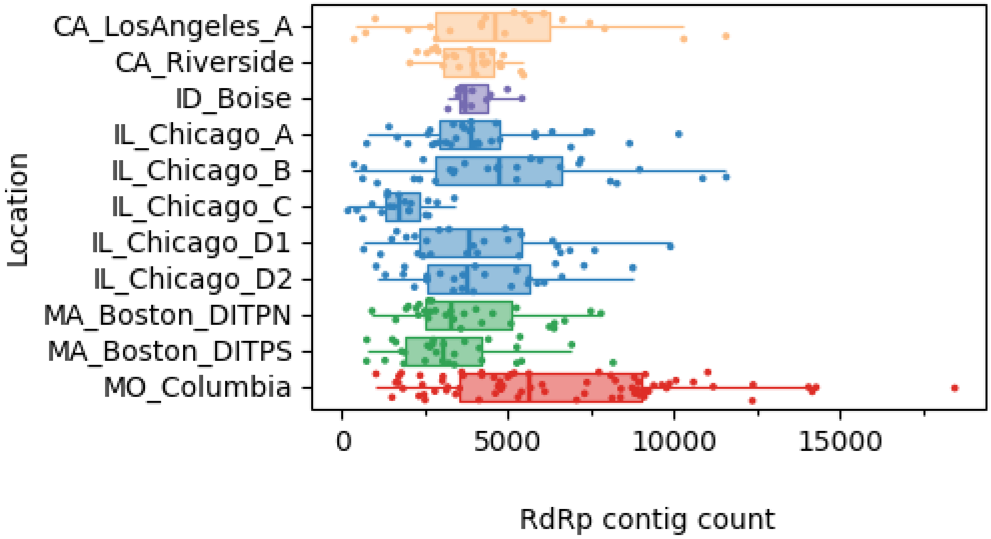

**B.**

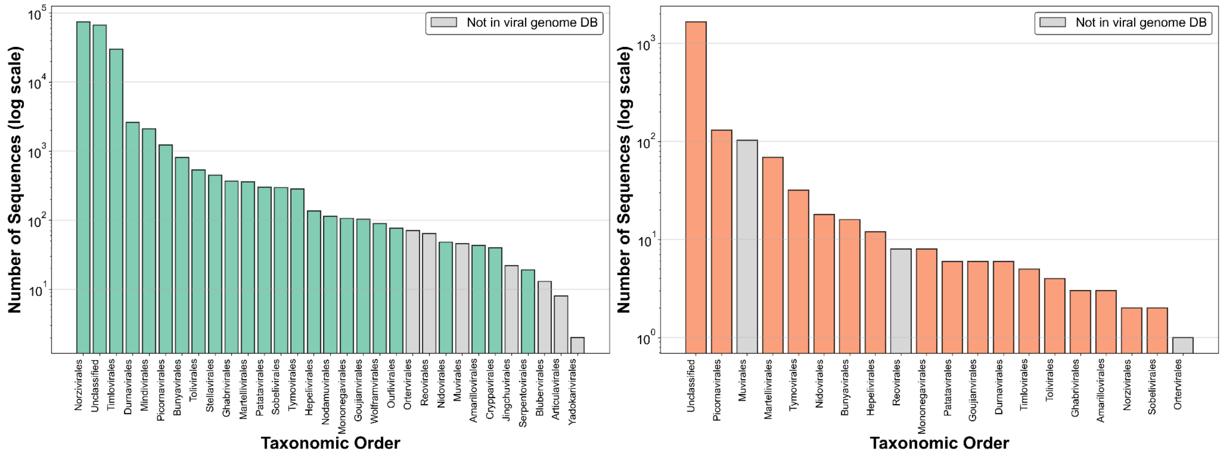


**Figure S3. (A)** Per-sample counts of assembled, near-complete RNA-dependent RNA polymerase genes. Samples are grouped by site. **(B)** Counts of RdRp clusters within each viral order. Clustering was performed at 95% identity and 75% alignment coverage. Clusters are categorized according to whether the RdRp protein contained the conserved motifs in the order "ABC" (left) or "CAB" (right).

**A.**

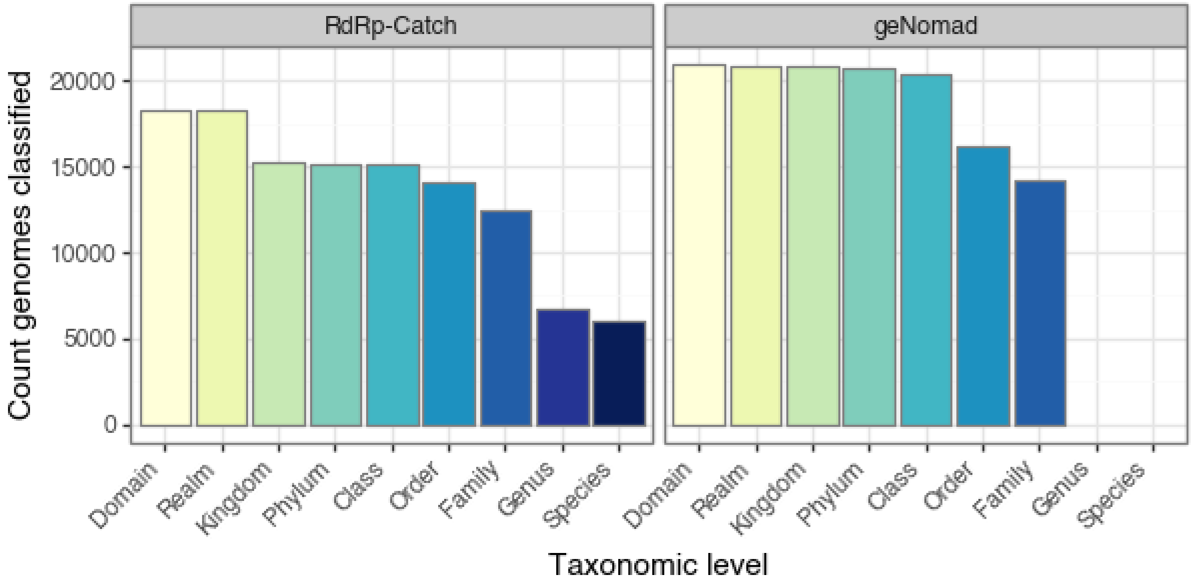


**B.**

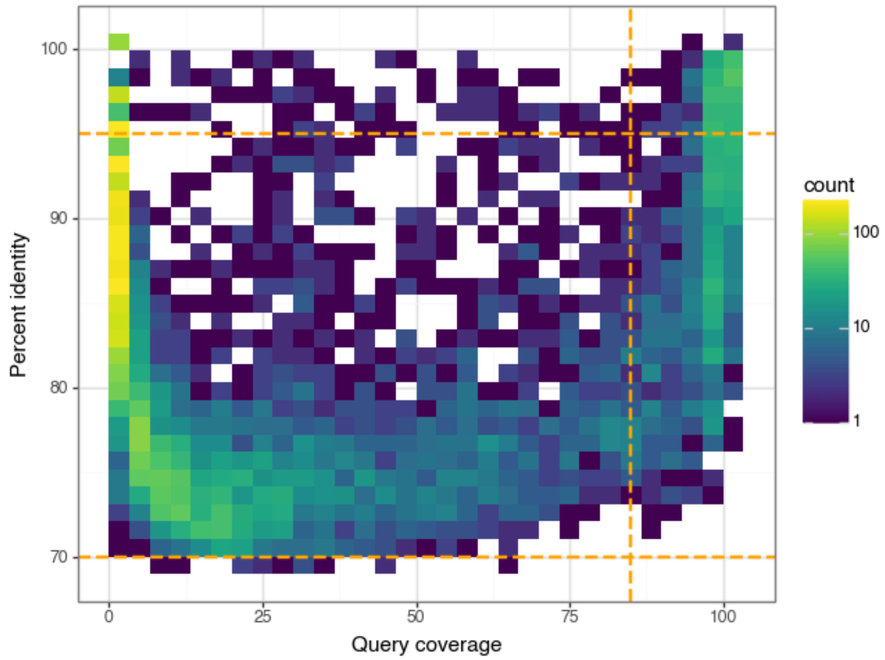


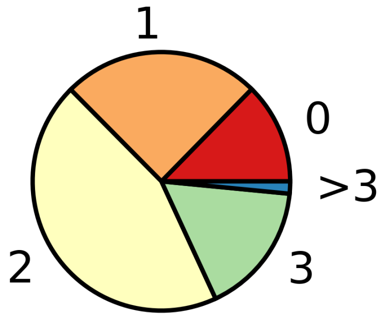


**Figure S4. (A)** Counts of genomes that could be classified down to each taxonomic level by two virus identification tools. Of note, geNomad is designed to classify RNA and DNA viruses (as low as viral family), while RdRp-CATCH is intended only for RNA viruses. **(B)** 2D density plot of results from BLASTn against NCBI core-nt with e-value < 1e-3 (July 2025), where multiple matches occurred, the hit with the highest product of query coverage and percent identity was included. The fill color of each cell indicates the count of genomes with best-hits at a given percent identity (y-axis) and query coverage (x-axis). Only 444 genomes had species-level hits (>95% identity and >85% query coverage, dashed orange lines) and 1453 had genus-level hits (>70% identity and >85% query coverage, dashed orange lines). Note log10 scaling of color. **(C)** Breakdown of vOTUs in WVDB based on the counts of VOGs from VOGDB per vOTU. Eighty-seven percent of vOTUs had at least one hit to VOGs in VOGDB.

**A.**

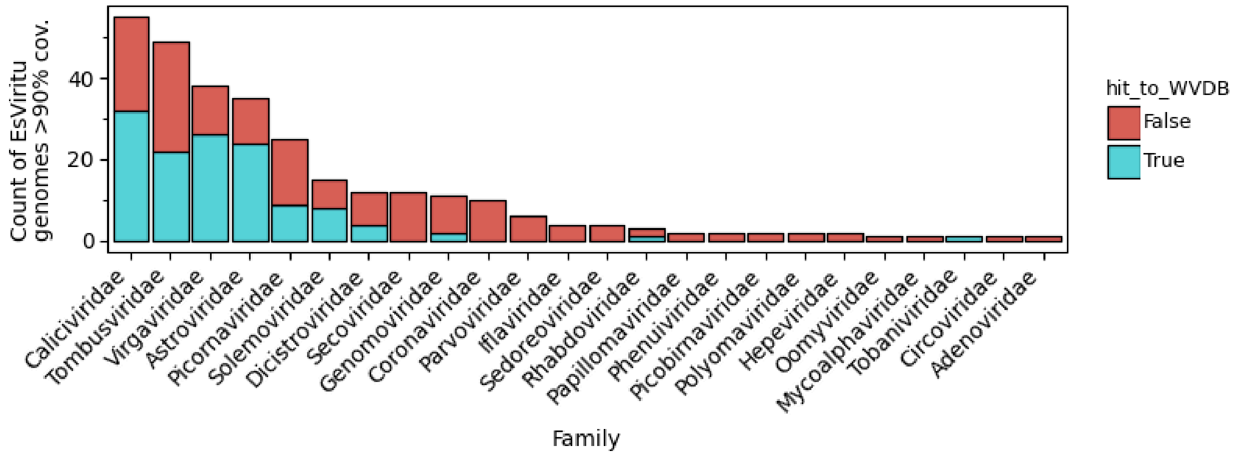


**B.**

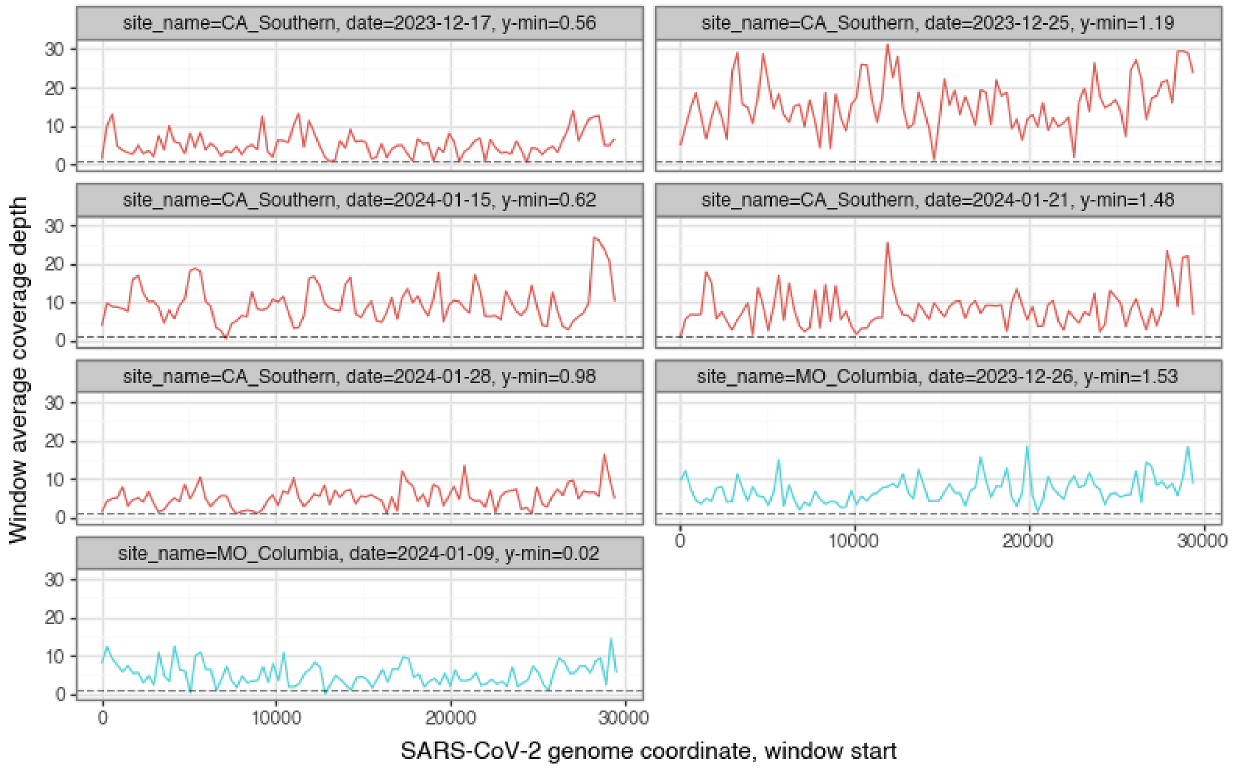


**Figure S5. (A)** Deduplicated count of virus reference sequences for which EsViritu reported a hit with >90% genome coverage in any of the 321 samples. Viruses are grouped by family level and colored by whether the EsViritu-generated consensus sequence had a species-level BLAST hit to a vOTU within WVDB. Where EsViritu reported hits to the same reference in multiple samples, the consensus sequence from the sample that produced the largest coverage breadth was chosen for comparison to WVDB. Secoviridae and Coronaviridae are notable families with multiple EsViritu hits but no representative in WVDB. **(B)** Sliding window average coverage depth information for SARS-CoV-2 reference genomes as generated by EsViritu. The 7 samples shown were selected because they contained ≥90% coverage breadth across a SARS-CoV-2 reference genome. However, no high-quality SARS-CoV-2 genomes were recovered from *de novo* assembly for inclusion in WVDB. In all samples, the average coverage for the window with the lowest coverage ("y-min" shown in facet labels) was around 1x (dashed line), suggesting a likely reason for discontiguous assemblies. When determining the minimum coverage, 500 nucleotides on the 5' and 3' ends of the genome were excluded.

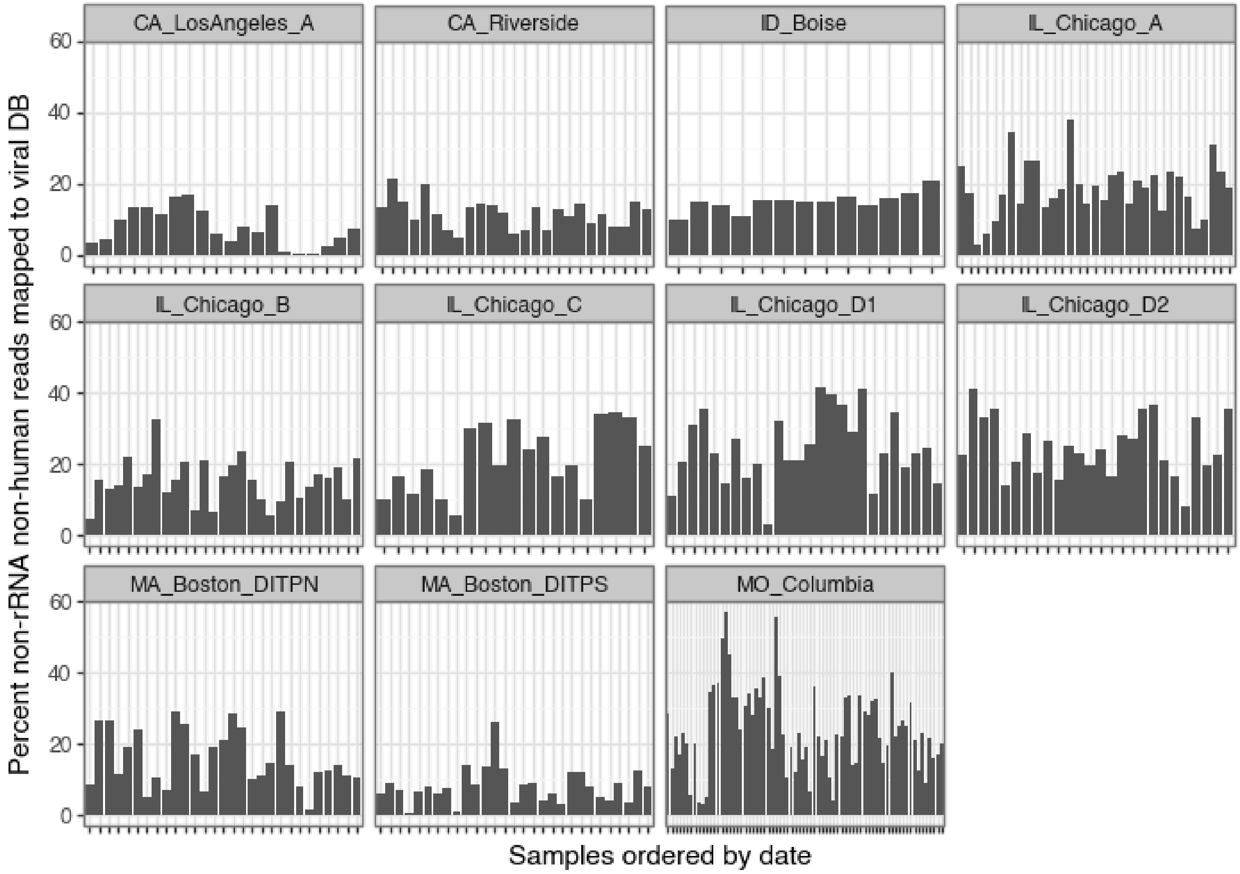


**Figure S6.** Percentage of reads mapped to the database across all sites and samples (independent x-axes).

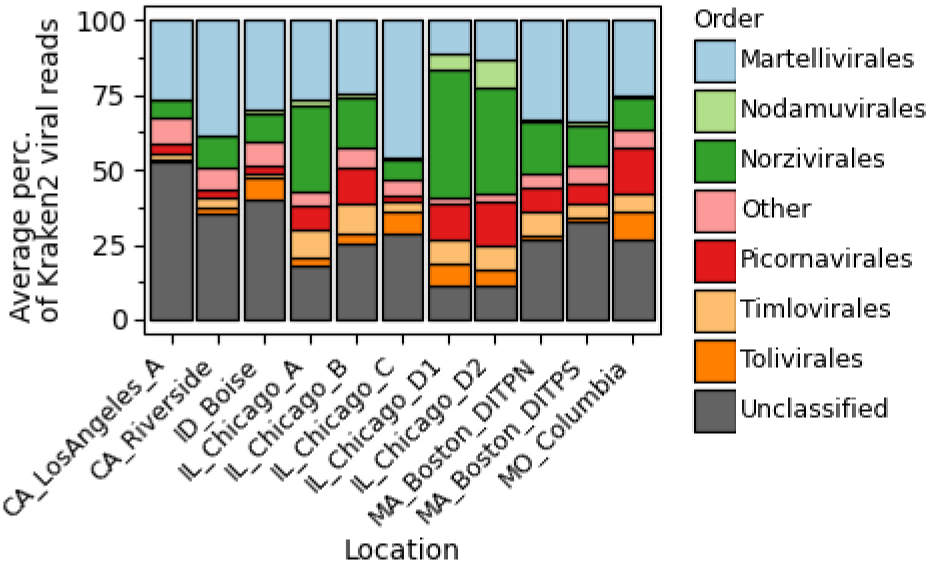


**Figure S7.** Order-level average relative abundance of only reads classified by Kraken2 with the NCBI core-nt database as viral. Here, 100% is the sum of the light and dark purple bar segments in Figure 2B. Relative abundances were averaged for each order across all samples per site. Orders whose maximum average relative abundance was below 5% are shown as "Other" (pink), while reads classified as viral but not given an order-level classification by Kraken2 are shown as "Unclassified" (gray). Note that while both Figure 3B and this figure show Order-level relative abundances, these figures are not directly comparable because they refer to different subsets of total reads.

**A.**

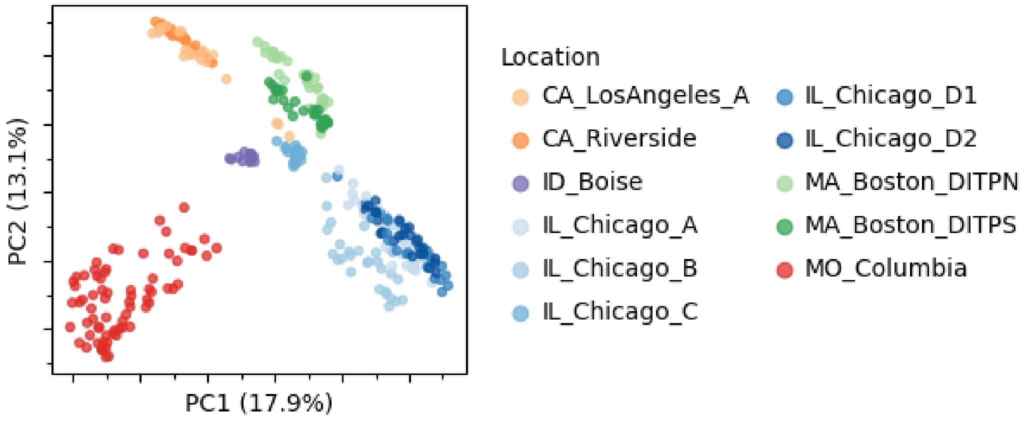


**B.**

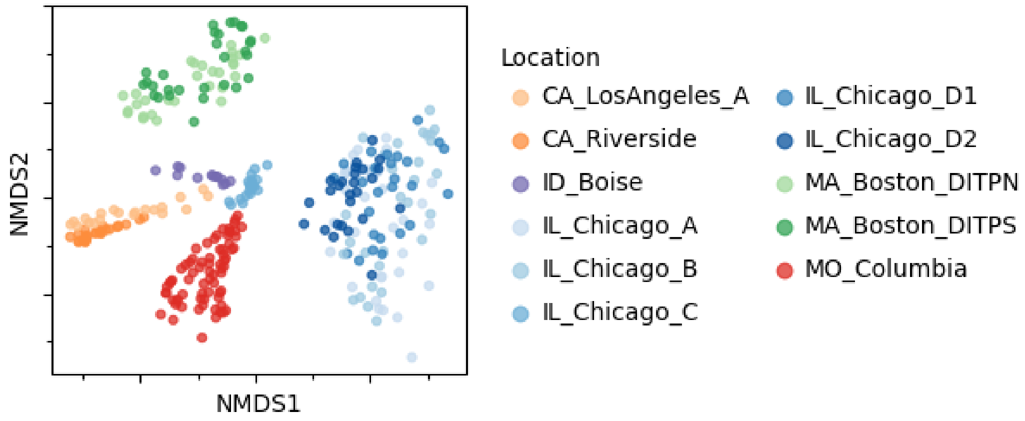


**C.**

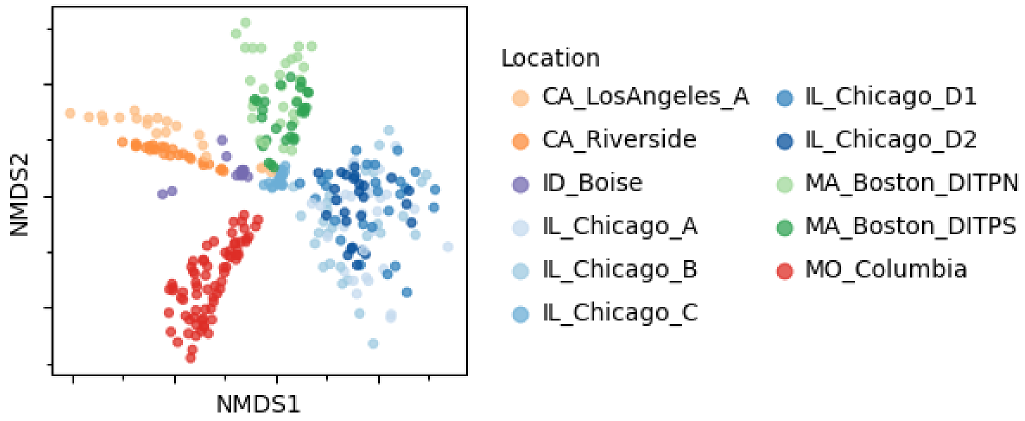


**Figure S8.** Additional ordination plots show similar patterns to the PCoA in Figure 3B: **(A)** PCoA made with read counts as abundance, **(B)** non-metric multidimensional scaling plot made with TPM as abundance, **(C)** non-metric multidimensional scaling plot made with read counts as abundance. All plots use Aitchison distance calculated from CLR-transformed abundance data. Points are colored by site and state as in Figure 3B.

**A.**

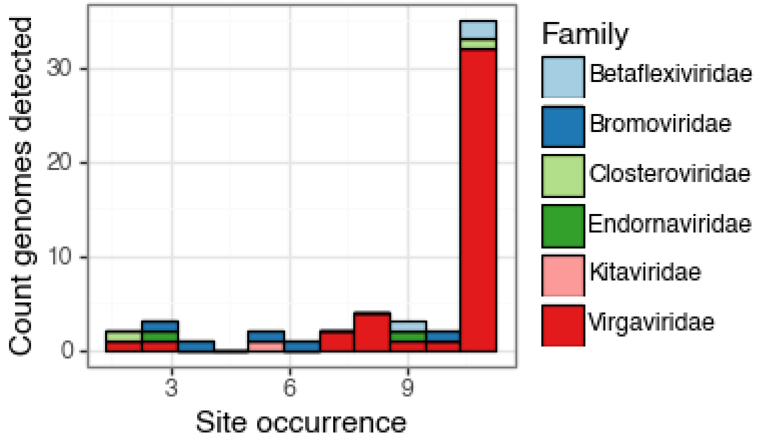


**B.**

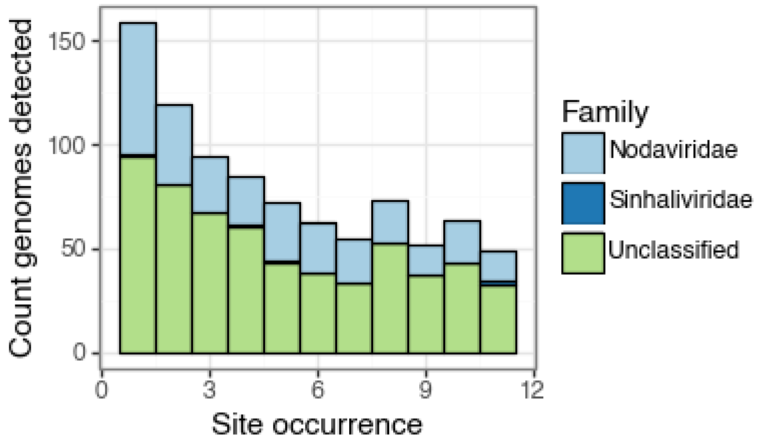


**C.**

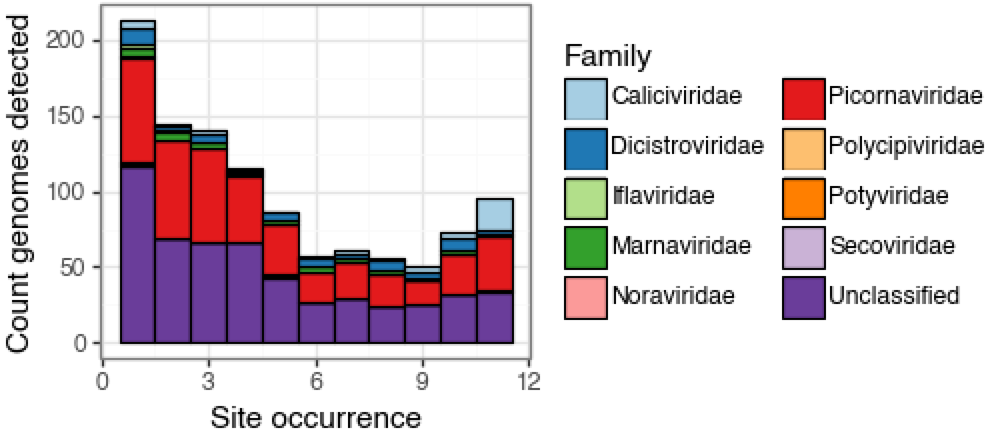


**Figure S9.** Counts of vOTUs (y-axis) categorized by the number of sites in which they were detected at least once (x-axis) for specific viral orders including (A) Martellivirales, (B) Nodavirales, and (C) Picornavirales. Counts are colored according to viral family within each order.
